# Temporal relationship between outbound traffic from Wuhan and the 2019 coronavirus disease (COVID-19) incidence in China

**DOI:** 10.1101/2020.03.15.20034199

**Authors:** Zaixing Shi, Ya Fang

## Abstract

**Background:** The city of Wuhan is the epicenter of the 2019 coronavirus disease (COVID-19) outbreak and a central Chinese hub for transportation and industry. Mass migration prior to the Chinese New Year may have accelerated the spread of COVID-19 across China. This analysis investigated the temporal relationship between daily outbound traffic from Wuhan and the incidence of COVID-19 in 31 Chinese provinces during January-February 2020.

**Methods:** We collected incidence of confirmed COVID-19 cases from National and Provincial Health Committee reports from January 10 to February 29, 2020. Volume of outbound traffic from Wuhan to other provinces was measured by Baidu Migration Index, a widely used metric that tracks migration based on cellphone location. We used cross-correlation function and autoregressive integrated moving average (ARIMA) model to examine time-lagged association between traffic volume and COVID-19 incidence by province. Contributors to the provincial variation in the temporal associations were investigated. Additionally, we estimated the reduction in cumulative incidence of COVID-19 cases following the travel ban for Wuhan.

**Results:** Cross-correlation function analyses suggested that the volume of outbound traffic from Wuhan was positively associated with COVID-19 incidence in all provinces, with correlation coefficients between 0.22-0.78 (all P<0.05). Approximately 42% of provinces showed <1 week of lag between traffic volume and COVID-19 incidence, 39% with 1 week, and 19% with 2-3 weeks. Migration had more prolonged impacts in provinces closer to Wuhan and with more passenger influx from Wuhan, but affected economically advantaged provinces to a lesser extent. We further estimated that the travel ban may have prevented approximately 19,768 COVID-19 cases (95% CI: 13,589, 25,946) outside of Wuhan by February 29, 2020.

**Conclusions:** Outflowing migration from Wuhan facilitated the COVID-19 transmission to other parts of China with varying time-lagged effects dependent on provincial characteristics. The travel ban led to a significant reduction in COVID-19 outside of Wuhan.

## INTRODUCTION

The 2019 coronavirus disease (COVID-19) outbreak occurred during the mass migration period prior to the Chinese New Year in the city of Wuhan, Hubei province, a central Chinese hub for transportation and industry. On January 23, a travel ban put the city of Wuhan under quarantine, all transportations into and out of the city were canceled, impacting more than nine million people. It is reported that more than five million people have left Wuhan in the days before the travel ban [1], which may have accelerated the spread of COVID-19 to other provinces in China. However, the dynamic relationship between outbound traffic from Wuhan and COVID-19 outbreak in other parts of China remains unclear.

Several analyses have evaluated the impact of migration on the COVID-19 outbreak. For example, Du et al. estimated that COVID-19 may have been transported from Wuhan to more than 130 Chinese cities and all four major metropolitan areas by January 23 [2]. Tian et al. reported that the travel ban slowed the transmission of infection to other Chinese cities by an average of 2.9 days [3]. Chinazzi et al. suggested that the travel ban delayed the overall epidemic progression of COVID-19 in China by 3-5 days, and may reduce the number of cases outside of China by 80% by the end of February [4]. Further, Jin et al. and Chen et al. reported that the cumulative migration from Wuhan was highly correlated with the number of reported cases in other provinces in China, with correlation coefficients ranging from 0.5 to 0.9 [5, 6]. However, no study to date has estimated the time lag between traffic and disease incidence and the total number of cases prevented by the travel ban.

This analysis aimed to fill in the gap by examining the time-lagged effect between the volume of outbound traffic from Wuhan with the COVID-19 incidence in 31 Chinese provinces during January-February 2020. We further evaluated the overall intervention effect of the Wuhan travel ban by the end of February. Our study used time-series analysis to estimate the association between travel volume and subsequent COVID-19 incidence, which is less biased as it accounts for the strong autocorrelation within data that are generated from sequential observations.

## METHODS

### Data

#### COVID-19 incidence data

We collected the incidence of confirmed COVID-19 cases from National and Provincial Health Committee reports from January 10 to February 29, 2020. These reports were obtained from official government websites and state-run media. Data were aggregated by date and province.

#### Traffic data

Traffic data were collected through the Baidu Migration Map (https://qianxi.baidu.com/), an online interactive map developed by Baidu Inc. to display human migration volume for Chinese cities. The map displays inbound and outbound traffic volumes for selected cities and dates. Traffic volume is expressed as the Baidu Migration Index, a proprietary index derived based on cell phone positioning data and is proportional to the daily total number of people traveling out of Wuhan. Although the explicit relationship between the population size and the Baidu Migration Index is unclear, previous analysis has estimated that one unit of the index corresponds to approximately 44,520 travelers [4]. Using web scraping technique, we obtained the Baidu Migration Index data for outbound traffic from Wuhan to the 31 Chinese provinces from January 10 to February 29, 2020. We also extracted the percentage of people who traveled to each province from Baidu Migration Map. With these data, we estimated the volume of daily traffic from Wuhan to each province, calculated as Volume_province,date_ = Total Volume_date_ × Percentage_province,date_. Data for the same Lunar calendar period in 2019 were used as benchmark to make predictions for disease incidence if the travel ban had not been issued.

#### Contextual data for provinces

We included demographic, economic, and geographical factors that are potential contributors to the provincial variation in the association between traffic volume and COVID-19 incidence. These factors included geographical distance between Wuhan and each province’s capital city, the average daily outbound traffic from Wuhan to each province, the gross domestic product (GDP) per capita, and the population density of each province. Geographical distances between cities were calculated based on their coordinates. Average traffic volumes were estimated based on the 2019 Baidu Migration Index. Economic and demographical data were from the Chinese Bureau of Statistics [7].

## Statistical analyses

We used time series analyses, including cross-correlation function (CCF) and autoregressive integrated moving average (ARIMA) model [8], to examine time-lagged association between traffic volume and COVID-19 incidence. Time series analysis is appropriate because it examines the correlation between two time series data and makes predictions while accounting for any autocorrelation structure within each time series and any shared trends. We used the CCF to understand the time-lagged correlation between traffic volume and COVID-19 incidence. A prewhitening process was applied to the time-series data to avoid common trends between traffic and CVID-19 incidence [9]. The prewhitening was conducted by first fitting an ARIMA model on the x-variable and calculating the residuals, and then filtering the y variable using the x-variable ARIMA model. The CCF assesses the correlation between the residuals from the x-variable ARIMA model and the filtered y values. In our case, the x variable is the daily traffic volume time series and the y variable is the COVID-19 case time series. The non-seasonal ARIMA model is generally denoted as ARIMA (*p, d, q*), in which *p* is the order of the autoregression (AR) component, *d* is the order of the differencing process to form a stationary times series, and *q* is the order of the moving average (MA) process. In an ARIMA model, the value of *y* at time *t* is estimated as:

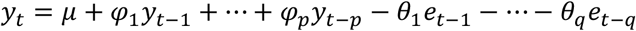

where *y*_*t*_ is the value at time *t, φ* is the AR parameter, and *θ* is the MA parameter.

Additionally, we evaluated the potential reduction in the cumulative incidence of COVID-19 cases in other parts of China following the Wuhan travel ban. To account for the variation in the association between traffic volume and COVID-19 incidence by province, we first classified provinces according to their estimated time lag and correlation coefficients using K-means clustering [10]. For each province cluster, we fitted an ARIMA model with traffic volume as an external regressor, also known as an ARIMAX model. The ARIMAX model describes the value of *y* at time *t* using the following equations:

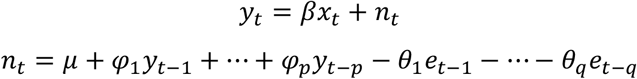

in which *x*_*t*_ is a covariate at time *t* and *β* is its coefficient. The lag values identified by the cross-correlation analysis was applied to traffic time series data in the ARIMAX model. The models were fitted using data up to the expected turning point in COVID-19 incidence following the travel ban. We used the median lag time for each cluster to determine the turning points. We used differencing to stationarize the COVID-19 incidence time series and checked stationarity using the augmented Dickey-Fuller test. The fitted ARIMAX model was used to forecast cumulative incidence till the end of February based on migration data during the same period in 2019. The difference between the predicted and observed cumulative incidence is the estimated intervention effect associated with the travel ban.

Statistical analyses were two-sided with type I error rate of 5%. We used R 3.6.1 for all analyses.

## RESULTS

Outbound traffic from Wuhan started to increase rapidly from January 13 and fell sharply after January 23, 2020, when the travel ban was implemented (Figure 1A-1C). By January 23, 71% of people who traveled out of Wuhan went to other cities in Hubei province and 29% migrated to other provinces. The incidence of COVID-19 within and outside of Hubei province fluctuated in similar patterns with the change in traffic (Figure 1E-1G), suggesting a lagged association between traffic volume and COVID-19 incidence.

**Figure 1.**
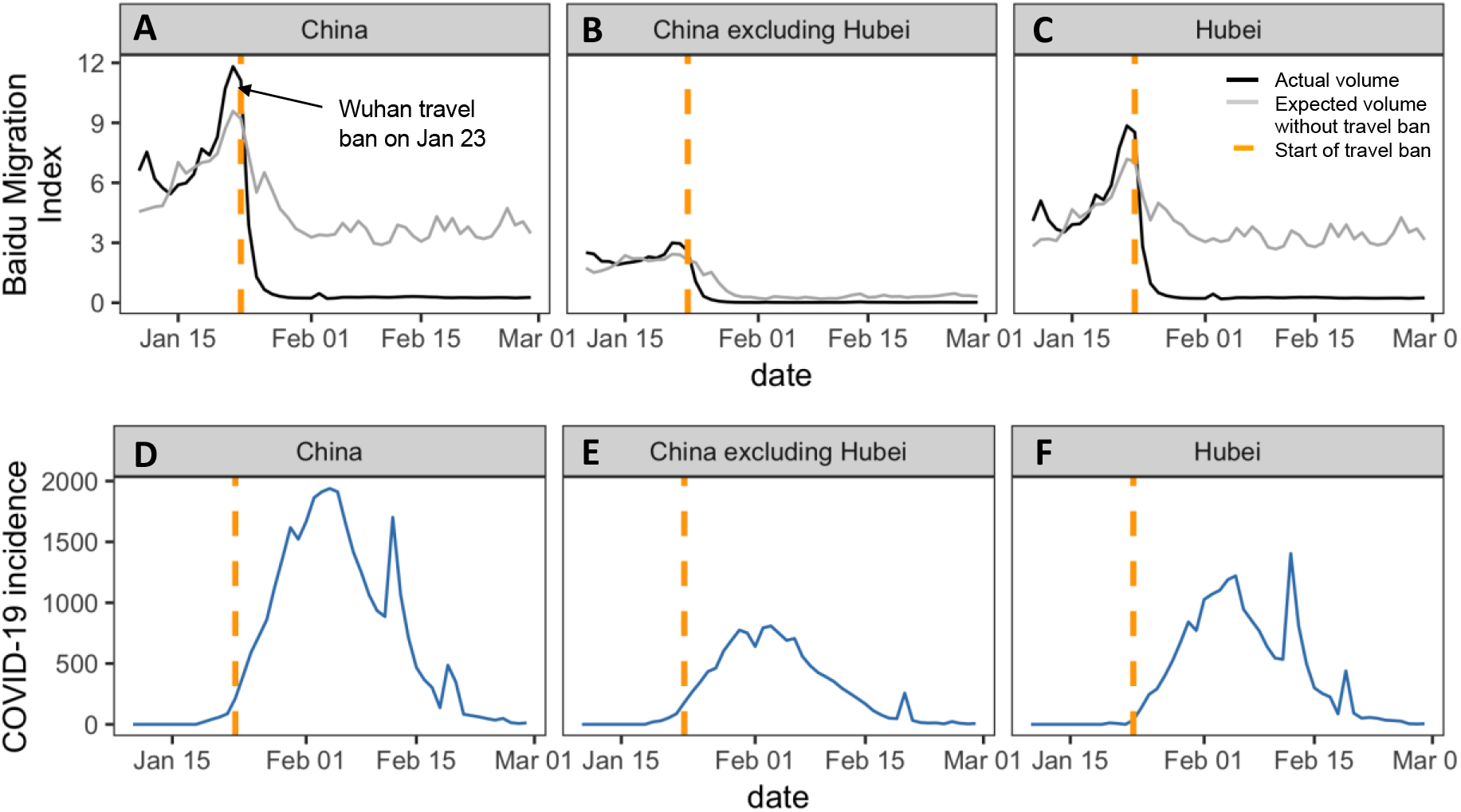
Trend of outbound traffic from Wuhan and incidence of COVID-19 in China. Fig. 1A-1C displays the Baidu Migration Index from January 10 to February 29. The black lines indicate actual traffic volume under the travel ban and the grey lines show the potential traffic volume without the travel ban. The yellow vertical line at January 23 shows when the Wuhan travel ban began. Fig. 1D-1F shows the trend of COVID-incidence from January 10 to February 29.

After prewhitening the COVID-19 incidence time series by the ARIMA model fitted on the outbound traffic time series, we calculated the cross-correlation coefficients between daily outbound traffic volume and COVID-19 incidence for each province. There were great geographical variations in time lags and correlation coefficients (Figure 2A-2B). K-means clustering analysis identified 3 latent clusters of provinces according to time lag and correlation coefficients (Figure 2C). The estimated time lags between traffic volume and COVID-19 incidence were <1 week in 42% of provinces, 1 week in 39% of provinces, and 2-3 weeks in 19% of provinces.

**Figure 2.**
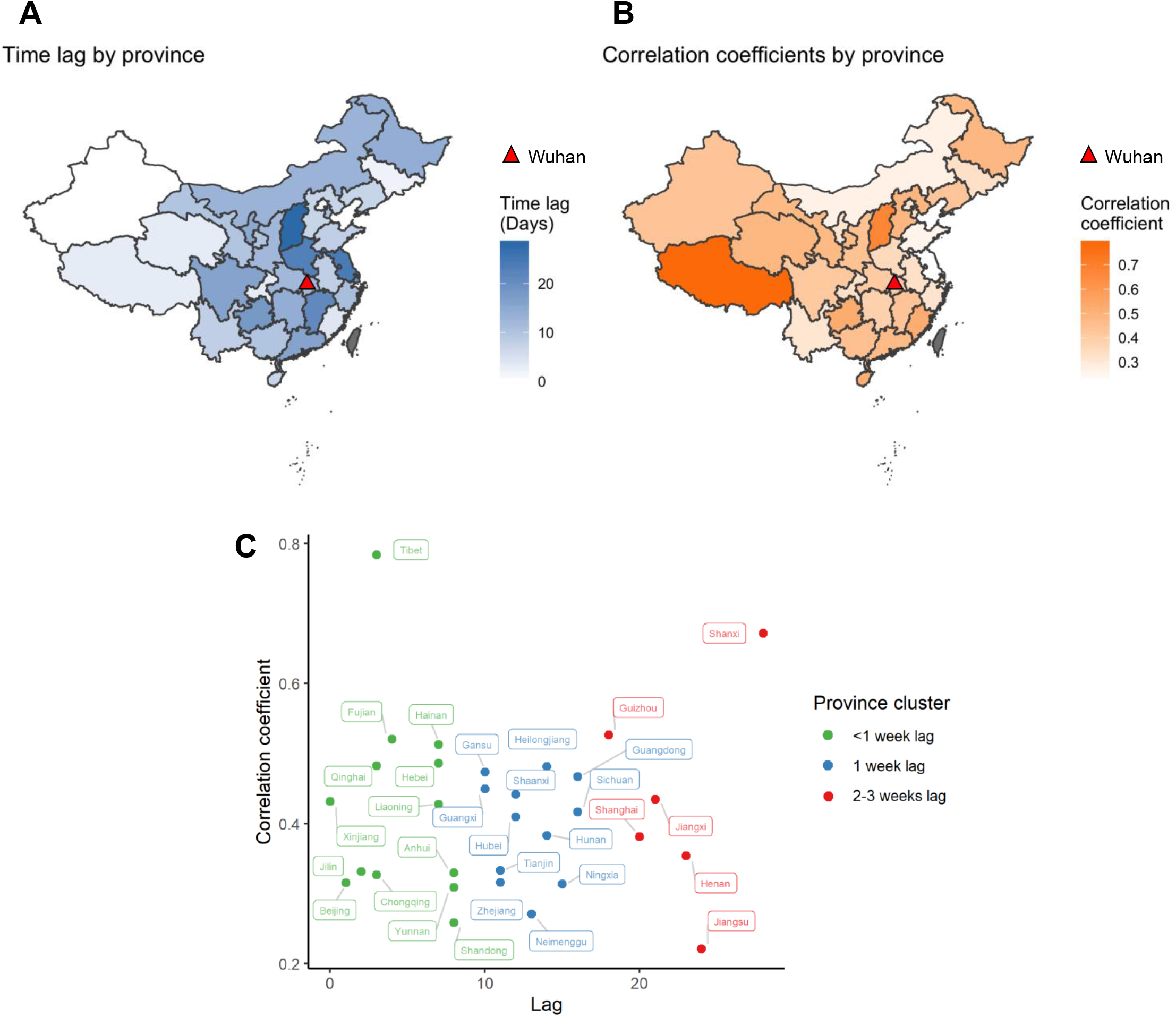
Provincial variations in time lags and correlation coefficients between volume of traffic from Wuhan and COVID-19 incidence. Fig. 2A shows the variation in time lag for the association between traffic from Wuhan and COVID-19 incidence by province, with darker color indicating shorter lag. The red triangle indicates the location of Wuhan. Fig. 2B shows the variation in correlation coefficients by province, with darker color indicating stronger correlation. Fig. 2C displays the 3 distinct clusters of provinces based on lags and correlation coefficients using K-means clustering.

We examined the potential contributors to provincial variations in the association between traffic volume and COVID-19 incidence. The analyses showed that the time lags of the migration effect were negatively associate with geographical distance (β= -4.9 per 1000 km, P=0.004; Figure 3A) and positively with average daily traffic volume (β= 86.9 per unit, P=0.02; Figure 3B). Correlation coefficients were positively associated with geographical distance (β=0.1 per 1000 km, P=0.03; Figure 3E) and negatively with GDP (β= -0.01 per 1000 USD/person, P=0.02; Figure 3G). Population density was not associated with either the lag or correlation coefficient. However, in sensitivity analyses excluding Shanghai from the analysis, whose population density far exceeds all other provinces, population density was negatively associated with the correlation coefficient (β= -0.15 per 1000 persons/sq.km, P=0.03).

**Figure 3.**
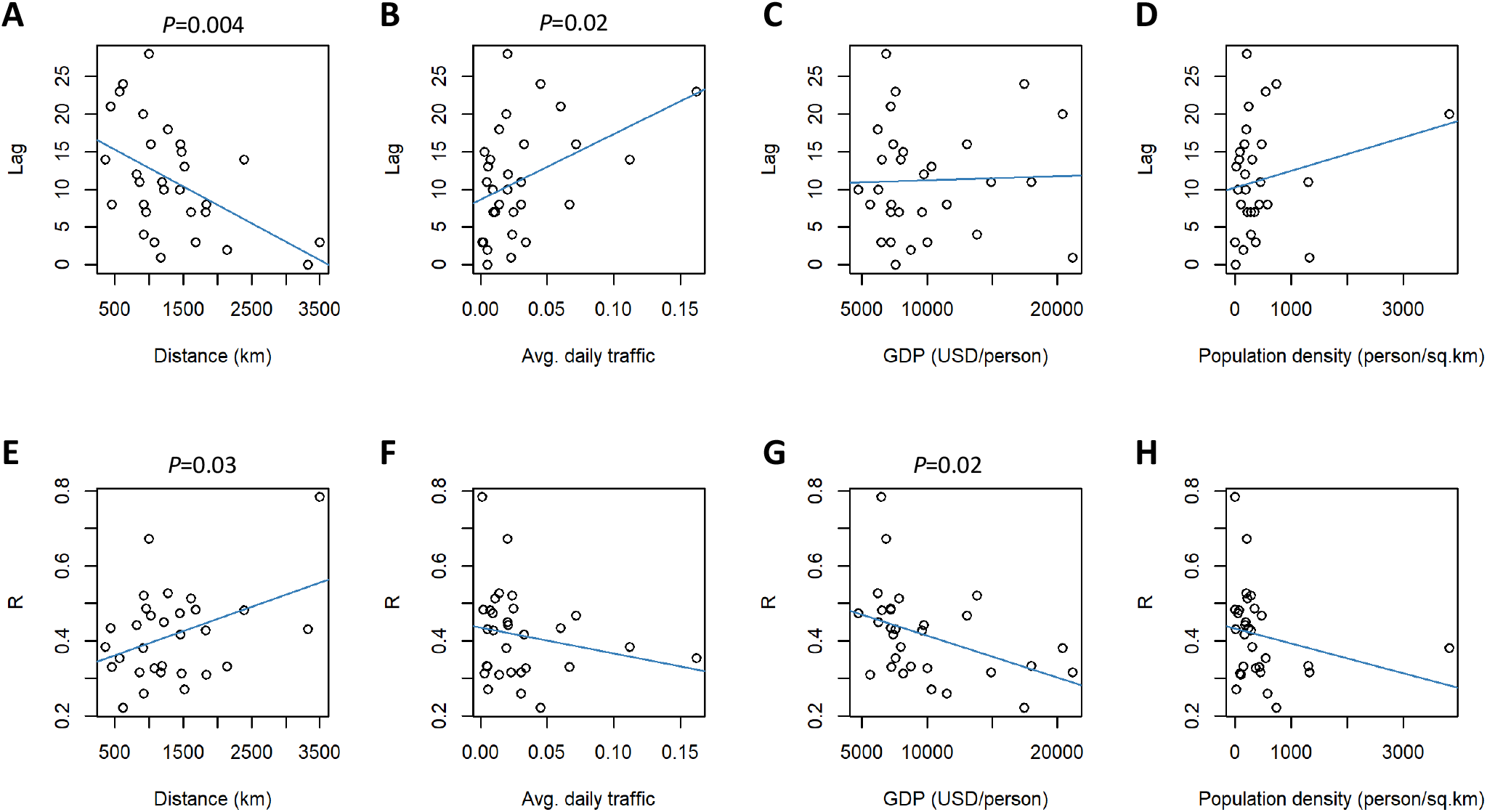
Relationship between time lags and correlation coefficients with distance to Wuhan, average daily traffic from Wuhan, GDP, and population density. Fig. 3A-3D shows the correlation of time lag with contextual factors, and Fig. 3E-3H shows the correlation between correlation coefficients and the contextual factors.

The optimal ARIMAX models for each province cluster are summarized in Table 1. The ARIMA (2,1,0) was optimal for provinces with <1 week of lag, and the ARIMA (1,1,0) was optimal for provinces with ≥1 week of lag. The model-predicted cumulative incidence based on actual traffic data demonstrated good fit with the observed incidences (Figure 4A-4C, Table 1). The predicted cumulative incidences would be higher in all three clusters of provinces based on 2019 traffic data (Figure 4D-4F), and the differences were projected to grow larger moving forward. It is estimated that there would be approximately 2,123 more cases in provinces with <1 week lag, 4,363 more in provinces with 1 week of lag, and 13,282 more in provinces with 2-3 weeks of lag by the end of February, although the predicted value was not significantly different from the observed in provinces with <1 week of lag (Table 1). We estimated that the travel ban may have reduced a total of 19,768 (95% CI: 13,589, 25,946) cases outside of Wuhan by the end of February.

**Table 1.** Model fitting for the ARIMA models and model predictions.

| Province cluster<br>by time-lagged<br>effect | Percent of<br>provinces | Model | RMSE | MAE | AIC | BIC | Predicted cumulative<br>incidence without travel<br>ban by Feb 29, 2020<br>(95% CI) | Observed<br>cumulative<br>incidence by<br>Feb 29, 2020 |
| --- | --- | --- | --- | --- | --- | --- | --- | --- |
| <1 week lag | 42% | ARIMA (2,1,0) | 25.48 | 21.33 | 135.86 | 140.59 | 7,405 (3,522, 11,288) | 5,282 |
| 1 week lag | 39% | ARIMA (1,1,0) | 23.68 | 18.23 | 102.35 | 107.23 | 10,046 (9,950, 10,141) | 5,683 |
| 2-3 weeks lag | 19% | ARIMA (1,1,0) | 159.21 | 136.33 | 191.14 | 197.12 | 33,413 (31,213, 35,613) | 20,131 |

Note:
Abbreviations: ARIMA, autoregressive integrated moving average; RMSE, root mean square error; MAE, mean absolute error; AIC, Akaike's Information Criteria; BIC, Bayesian Information Criteria; 95% CI, 95% confidence interval

**Figure 4.**
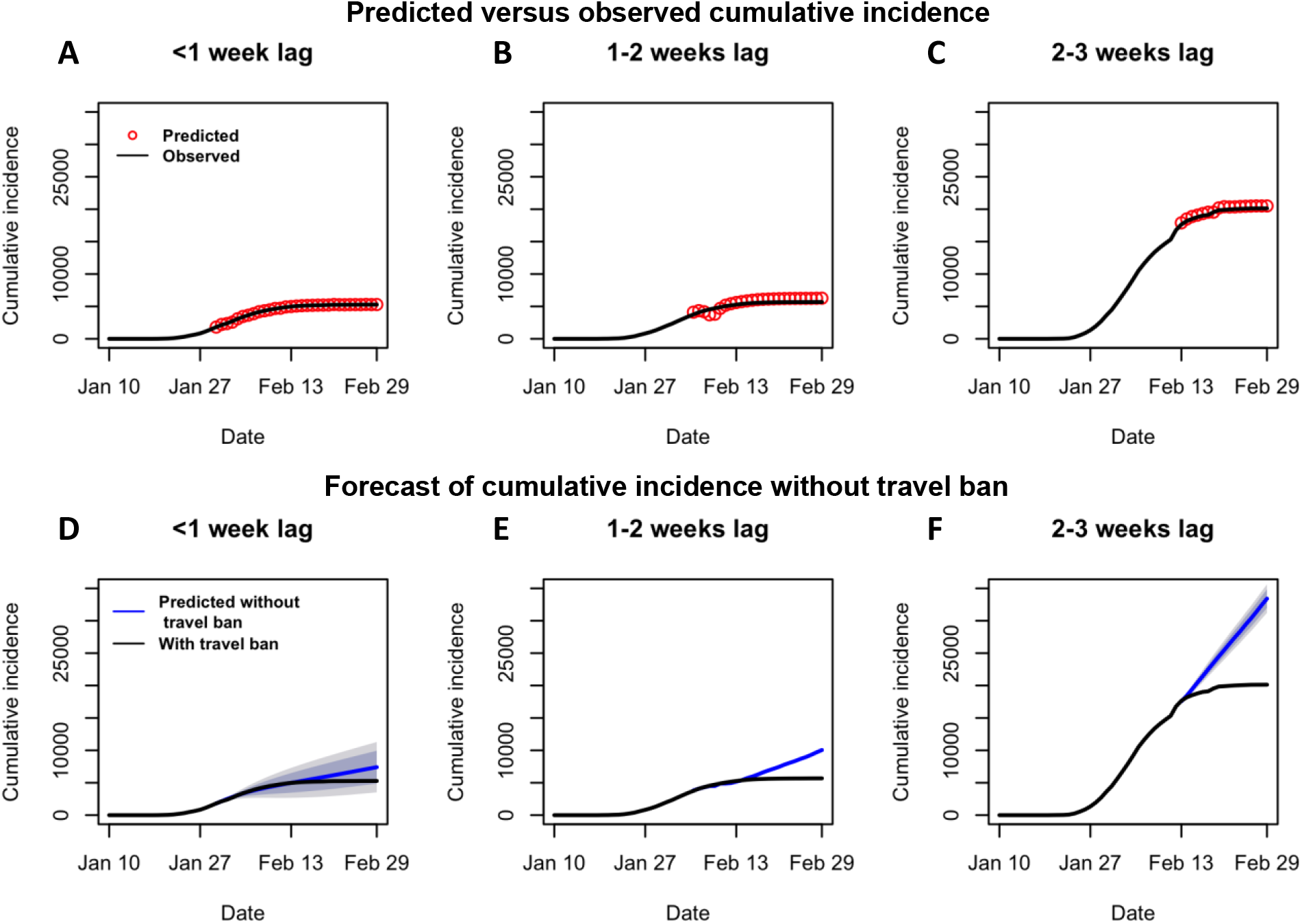
Predicted cumulative incidence of COVID-19 based on actual and historical traffic data. Fig. 4A-4C show that the predicted incidence were well fitted with observed data. Red dots are predicted incidence based on actual traffic data in 2020, and the black lines are observed incidence. Fig. 4D-4F show that the cumulative incidences were projected to increase had the travel ban not been implemented. The blue lines are predicted incidence based on historical travel data for the same Lunar calendar period in 2019, and the black lines are the observed incidence.

## DISCUSSION

In this analysis, we examined the temporal relationship between outbound traffic from Wuhan and the incidence of COVID-19 in 31 Chinese provinces. Our analyses suggest a positive time-lagged association between human migration and the incidence of COVID-19. We identified 3 distinct clusters of provinces based on the timing and strength of associations: 42% of provinces showed <1 week of lagged association, 39% with 1 week, and 19% with 2-3 weeks. Provinces that are closer to Wuhan and have more passenger influx from Wuhan experienced a more prolonged impact from migration. The correlations between traffic volume and COVID-19 incidence were weaker in economically advantaged provinces. Furthermore, we estimated that the travel ban for Wuhan may have prevented approximately 19,768 COVID-19 cases (95% CI: 13,589, 25,946) outside of Wuhan by the end of February. This is equivalent to a 39% reduction in expected cumulative incidence, suggesting that transportation control is an effective intervention to contain the COVID-19 outbreak in the absence of cure and immunization.

Regional variations in the association between outflowing traffic from Wuhan and COVID-19 incidence worth closer examination. Longer time lag between traffic volume and COVID-19 incidence was observed in provinces closer to Wuhan or with more population influx from Wuhan, such as Henan, Jiangxi, Shanghai, Guizhou, Guangdong, and Sichuan. Longer lagged effect does not mean these provinces were less responsive to imported cases. Rather, it means the overall changes in cumulative incidence in these provinces were not only driven by imported cases but also infected cases through local transmissions. For example, the lag was estimated to be 23 days for Henan province, which was severely affected by COVID-19. Although Henan province was among the earliest to implement traffic control, the COVID-19 incidences grew steadily after the traffic control. It is possible that the influx of Wuhan emigrants, mostly people who returned home from work/school, led to significant local transmissions in Henan, which outweighed the imported cases in driving the cumulative incidence to grow for a prolonged period. Given the median incubation period of 4-6 days for COVID-19 [11], the overall COVID-19 incidence in provinces with lags of 2-3 weeks, or approximately 2-3 times the median incubation period, could be reflective of the second- or third-generation transmissions. Also, provinces with longer lag tend to have higher cumulative incidence, suggesting widespread local transmissions.

Another possible reason for the prolonged impact of migration in provinces adjacent to Hubei is that cross-province migrations are hard to shut off given the geographical proximity. Sporadic travelers from Wuhan may continue to cross provincial borders and spread the virus for a while after the travel ban. This is evident by the fact that the lag was much shorter in provinces further away from Wuhan (e.g., Tibet, Qinghai, Xinjiang, Hainan, Yunnan) or had more strict traffic control (e.g., Beijing). Furthermore, travelers to these provinces were more likely to be tourists as opposed to returning home; therefore, they have little social ties to the local population and are less likely to introduce widespread local transmissions.

In addition, we found that the correlation between migration and COVID-19 incidence was lower in provinces with higher GDP per capita. For example, the correlation coefficients were between 0.2-0.4 for Beijing, Shanghai, Tianjin, and Jiangsu, provinces with GDP per capita over $15,000 US dollars in 2019. In contrast, the correlation coefficients were between 0.5-0.7 for Gansu, Guizhou, Heilongjiang, and Shanxi, provinces with GDP per capita below $7000 US dollars. The finding suggests that provinces with better healthcare resources, greater disease prevention efforts, and more mature infrastructure may have more capacity to manage disease outbreaks and maintain faster responses as the outbreak evolve.

The primary strength of this analysis is the use of ARIMA models to examine the temporal relationship between traffic and COVID-19 incidence, allowing us to disentangle the association from a myriad of shared common trends between the two variables. The analysis is also novel in that real-time, cellphone location-based migration data were used to measure population mobility, which is superior to methods based on historical mobility data. This analysis also fitted separate ARIMA models according to differing time-lagged impacts from traffic, leading to a more accurate estimate of the intervention effect for the travel ban.

This analysis is subject to limitations. First, changes in diagnostic capacity and criteria for COVID-19 may have contributed to, at least partly, the secular trend of COVID-19 incidence. For example, there were underdiagnoses of COVID-19 during the early stage of the outbreak, and later a surge in COVID-19 cases around February 13 due to broadened diagnostic criteria. These changes were not travel-related and may distort the estimated association to some extent. Second, the temporal association between traffic and COVID-19 could be confounded by other preventive measures, such as increased self-quarantine and widespread use of face mask and hand sanitization. Therefore, the estimated intervention effect associated with the travel ban could be overestimated. Finally, as an ecologic study, our findings may suffer from ecologic fallacy and do not necessarily imply individual-level association between travel history and risk of COVID-19 infection.

In conclusion, mass migration out of Wuhan facilitated the COVID-19 outbreak in other parts of China. The outflowing migration from Wuhan had a more prolonged impact on provinces that are closer to Wuhan and have more immigration from Wuhan. Provinces with higher GDP per capita were less impacted by migration. It is estimated that approximately 39% of cases outside of Wuhan have been prevented by the travel ban issued on January 23, 2020. In the absence of cures and immunization for an emerging infectious disease like COVID-19, transportation control, an old-fashioned approach, is proven an effective public health intervention. Our findings provide important evidence for government agencies to determine the optimal disease control strategy and improve preparedness for future public health emergencies.

## Data Availability

The data are available upon request to the authors.

## Notes

**Funding information:** This work was supported by the National Natural Science Foundation of China under grant number 81973144.

### Competing Interest Statement

The authors have declared no competing interest.

### Funding Statement

This work was supported by the National Natural Science Foundation of China under grant number 81973144.

